# Liver-specific polygenic risk score is more strongly associated than genome-wide score with Alzheimer’s disease diagnosis in a case-control analysis

**DOI:** 10.1101/2021.04.29.21256279

**Authors:** Daniel J. Panyard, Yuetiva K. Deming, Burcu F. Darst, Carol A. Van Hulle, Kaj Blennow, Gwendlyn Kollmorgen, Ivonne Suridjan, Cynthia M. Carlsson, Sterling C. Johnson, Sanjay Asthana, Corinne D. Engelman, Qiongshi Lu

## Abstract

Although our understanding of Alzheimer’s disease (AD) has greatly improved in recent years, the root cause remains unclear, making it difficult to find effective diagnosis and treatment options. Our understanding of the pathophysiology underlying AD has benefited from genomic analyses, including those that leverage polygenic risk score (PRS) models of disease. In many aspects of genomic research the use of functional annotation has been able to improve the power of genomic models. Here, we leveraged genomic functional annotations to build tissue-specific PRS models for 13 tissues and applied the scores to two longitudinal cohort studies of AD. The PRS model that was most predictive of AD diagnosis relative to cognitively unimpaired participants was the liver tissue score: n = 1,116; odds ratio (OR) (95% confidence interval [CI]) = 2.19 (1.70-2.82) per standard deviation (SD) increase in PRS; P = 1.46 × 10^−9^. After removing the *APOE* locus from the PRS models, the liver score was the only PRS to remain statistically significantly associated with AD diagnosis after multiple testing correction, although the effect was weaker: OR (95% CI) = 1.55 (1.19-2.02) per SD increase in PRS; P = 0.0012. In follow-up analysis, the liver PRS was statistically significantly associated with levels of amyloid (P = 3.53 × 10^−6^) and tau (P = 1.45 × 10^−5^) in the cerebrospinal fluid (CSF) (when the *APOE* locus was included) and nominally associated with CSF soluble TREM2 levels (P = 0.042) (when the *APOE* locus was excluded). These findings provide further evidence of the role of the liver-functional genome in AD and the benefits of incorporating functional annotation into genomic research.

## Introduction

Despite advances in our understanding of Alzheimer’s disease (AD), the causal mechanisms are not fully understood. Since the hallmarks of AD have long been its neuropathological findings[1], the brain has historically been the primary focus of investigations into AD etiology. However, recent research has implicated other systems in AD, including the immune system and processes of inflammation[2], the cardiovascular system[3], and the liver and metabolism[4]. The lack of clarity over the roles of these mechanisms and risk factors in AD hampers our ability to identify novel therapeutic targets and ultimately effective treatments[5].

Genomic research has provided a valuable tool for understanding upstream risk factors in AD. The importance of amyloid-β (Aβ), long known to aggregate and subsequently accumulate in plaques in the brain, has been underscored by the knowledge that familial AD can be driven by genetic mutations in genes directly impacting Aβ processing, like *APP, PSEN1*, and *PSEN2*[6]. In late-onset AD, genomic studies have repeatedly identified risk factors in a number of genes, like *APOE, CR1*, and *ABCA7*, expanding our knowledge of the molecular systems likely to be contributing to the development of AD[7–9]. For instance, the discovery of *TREM2* as a genetic risk loci is now being expounded in follow-up experimental research on soluble TREM2 levels, highlighting the role of microglia in AD[10–12].

One particular application of genomic research that can help tease apart the mechanisms contributing to a disease involves polygenic risk scores (PRS), which are a measure of risk composed of the contributions of many single nucleotide polymorphisms (SNPs). Genome-wide PRS models and related methods have already been useful in the study of AD, including predicting AD and age of onset[13–15], examining genetic risk beyond the *APOE* locus[16], identifying PRS-environment associations[17,18], and comparing the genetic basis of related forms of AD[19]. PRS models can further be enhanced by the incorporation of genome functional annotation, which can boost the prediction accuracy of a PRS for disease risk[17,20,21]. One of the benefits of incorporating functional annotation is that it can introduce information about tissues and cell types and their potential relevance to the genomics underlying a particular trait. In AD, where there is growing genetic evidence for the role of different tissue and cell types[22,23], such annotation can provide important information about the full spectrum of biology involved in AD.

Here, we leveraged cell-type-specific genomic functional annotations derived from epigenetic data [23–25] to create tissue-specific PRS models for AD. We estimated tissue-specific genetic risk scores for participants in two longitudinal cohort studies of AD and analyzed the association between each tissue-specific PRS and AD diagnosis. Further, given that AD pathophysiology like brain amyloidosis and tau pathology can be identified using cerebrospinal fluid (CSF) biomarkers, we examined the possible associations between these tissue-specific PRS models and different pathologies in AD using CSF biomarkers of neurodegeneration and inflammation.

## Materials and methods

### Study participants

Data from two longitudinal AD cohorts focusing on middle and older aged adults were used for this study. The first was the Wisconsin Registry for Alzheimer’s Prevention (WRAP) study, described previously[26]. Briefly, participants were between the ages of 40 and 65, fluent in English, able to perform neuropsychological testing, without a diagnosis or evidence of dementia at baseline, and without any health conditions that might prevent participation in the study.

The second cohort was the Wisconsin Alzheimer’s Disease Research Center (WADRC) study, described previously[27]. WADRC participants were categorized into one of six subgroups: 1) mild late-onset AD; 2) mild cognitive impairment (MCI); 3) age-matched healthy older controls (age > 65); 4) middle-aged adults with a positive parental history of AD; 5) middle-aged adults with a negative parental history of AD; and 6) middle-aged adults with indeterminate parental history of AD. The clinical diagnoses for these groups were based on the National Institute of Neurological and Communicative Disorders and Stroke and Alzheimer’s Disease and Related Disorders Association (NINCDS-ADRDA)[28] and National Institute on Aging and Alzheimer’s Association (NIA-AA)[29]. Briefly, participants were over the age of 45, able to fast from food and drink for 12 hours, with decisional capacity, and without a history of certain medical conditions (like congestive heart failure or major neurologic disorders other than dementia) or any contraindication to biomarker procedures.

This study was performed as part of the GeneRations Of WRAP (GROW) study, which was approved by the University of Wisconsin Health Sciences Institutional Review Board. Participants in the WADRC and WRAP studies provided written informed consent. STREGA reporting guidelines[30] were used in the description of the results.

### Clinical diagnoses

AD, MCI, and other diagnoses of cognitive status for both WRAP and WADRC were made by a consensus review committee comprising an expert panel of dementia-specialist physicians, neuropsychologists, and nurse practitioners[26]. CSF biomarker status was not used in the process of making the clinical diagnoses.

### CSF biomarkers

Details on the collection of CSF for biomarker analyses have been previously described[31]. Briefly, CSF samples were acquired using a uniform preanalytical protocol between 2010 and 2018. Samples were collected in the morning using a Sprotte 24- or 25-gauge atraumatic spinal needle, and 22 mL of fluid was collected via gentle extraction into polypropylene syringes and combined into a single 30 mL polypropylene tube. After gentle mixing, samples were centrifuged to remove red blood cells and other debris. Then, 0.5 mL CSF was aliquoted into 1.5 mL polypropylene tubes and stored at -80 degrees Celsius within 30 minutes of collection.

All CSF samples were assayed between March 2019 and January 2020 at the Clinical Neurochemistry Laboratory at the University of Gothenburg. CSF biomarkers were assayed using the NeuroToolKit (Roche Diagnostics International Ltd., Rotkreuz, Switzerland), a panel of automated robust prototype immunoassays designed to generate reliable biomarker data that can be compared across cohorts. Measurements with the following immunoassays were performed on a cobas e 601 analyzer: Elecsys® β-amyloid (1–42) CSF (Aβ42), Elecsys Phospho-Tau (181P) CSF, and Elecsys Total-Tau CSF, β-amyloid (1–40) CSF (Aβ40), and interleukin-6 (IL-6). The remaining NTK panel was assayed on a cobas e 411 analyzer, including markers of synaptic damage and neuronal degeneration (neurogranin, neurofilament light protein [NFL], and α-synuclein) and markers of glial activation (chitinase-3-like protein 1 [YKL-40] and soluble triggering receptor expressed on myeloid cells 2 [sTREM2]).

A total of 9 CSF biomarkers were analyzed in this study: the Aβ42/Aβ40 ratio, phosphorylated tau 181 (ptau), the ptau/Aβ42 ratio, NFL, α-synuclein, neurogranin, YKL-40, sTREM2, and IL-6. Since the CSF biomarker measurements were to be used as outcomes, each biomarker was assessed for skewness using the skewness function of the R package moments (version 0.14) [32]. Any biomarker with a skewness ≥ 2 was transformed with a log_10_-transformation to better meet the normality assumption of regression. The outcomes that were log_10_-transformed were ptau, the ptau/Aβ42 ratio, NFL, and IL-6.

### Genomic data

The genomic data collection in the WRAP and WADRC cohorts has been described previously[33]. The WRAP samples were genotyped using DNA from whole blood samples and the Illumina Multi-Ethnic Genotyping Array at the University of Wisconsin Biotechnology Center[34]. Samples and variants with high missingness (> 5%) or inconsistent genetic and self-reported sex were removed. Samples from individuals of European descent were then imputed using the Michigan Imputation Server[35] and the Haplotype Reference Consortium (HRC) reference panel[36], with low quality variants again removed (R^2^ < 0.8) post-imputation. A total of 1,198 samples with 10,499,994 SNPs were present at the end of quality control.

Whole blood samples from the WADRC were genotyped by the Alzheimer’s Disease Genetics Consortium (ADGC) at the National Alzheimer’s Coordinating Center (NACC) using the Illumina HumanOmniExpress-12v1_A, Infinium HumanOmniExpressExome-8 v1-2a, or Infinium Global Screening Array v1-0 (GSAMD-24v1-0_20011747_A1) BeadChip assay. Each chip’s genomic data were initially processed separately. After strict quality control that removed variants or samples with high missingness (> 2%), out of Hardy-Weinberg equilibrium (HWE) (P < 1×10^−6^), or with inconsistent genetic and self-reported sex, samples were imputed with the Michigan Imputation Server where they were phased using Eagle2[37] and imputed to the HRC reference panel. Low quality variants (R^2^ < 0.8) or out of HWE were removed. After imputation, the data sets from the different chips were merged together, leaving a data set with 377 samples of European descent and 7,049,703 SNPs.

To prevent variant overlap issues, all genomic data sets used in this study were harmonized, including the WADRC, WRAP, International Genomics of Alzheimer’s Project (IGAP) 2019 GWAS of AD[7], and the 1000 Genomes Utah residents with Northern and Western European ancestry (CEU)[38] data sets. The GRCh37 genome build was used, all ambiguous SNPs were removed, and all SNPs were aligned to have strand and allele orientations consistent with the WADRC data set. Only the subset of SNPs successfully harmonized and present across all four data sets were used to build the PRS. To avoid sample overlap between participants in the WADRC and IGAP, 165 overlapping participants were removed, leaving a total of 1,410 participants (212 WADRC, 1,198 WRAP) with 5,631,405 SNPs.

### Tissue-specific functional SNP sets

To categorize the genome into tissue-specific functional regions, annotations from GenoSkyline-PLUS[23–25] (v 1.0.0) were used. GenoSkyline is an unsupervised framework that uses epigenetic data sets from the Roadmap Epigenomics Project[39] to predict tissue-specific functional regions of the genome. For each region of the genome and for each cell type, GenoSkyline predicts a value between 0 and 1 that represents whether that region is likely to be functional for that cell type, with 1 indicating functional and 0 non-functional. Due to the bimodal distribution of these scores, values ≥ 0.5 were considered functional and values < 0.5 were considered non-functional in this study. GenoSkyline-PLUS annotations are available for a variety of cell types, with each cell type labeled as part of a larger tissue type. The full list of GenoSkyline-PLUS tissues and included cell types can be found in Supplementary Table 1. For each tissue, the union of all functional genomic regions from all included cell types was defined as the functional genomic region for that tissue, and all SNPs falling within that region were included in that tissue’s set of tissue-specific functional SNPs. A total of 13 tissue-specific SNP sets were defined in this manner, with a 14^th^ SNP set comprising all SNPs regardless of tissue functionality to create a non-tissue-specific genome-wide PRS for comparison. For all of the PRS models, the effect size estimate for each SNP came from the 2019 IGAP GWAS results[7]. The nearest protein-coding gene to each SNP was also added using GENCODE (version 19) annotations[40].

Since certain genotypes in the *APOE* locus are known to be strongly associated with AD, we sought to examine whether the PRS were associated with AD beyond the effect of the *APOE* locus. To do so, we built tissue-specific PRS using the same procedure as above with the exception that all SNPs in a window around the *APOE* genomic region were removed (defined as between the *PVR* and *GEMIN7* genes: chromosome 19, base pairs 45,147,098-45,594,782). Some SNPs from the *APOE* locus were considered functional for each tissue, so the PRS scores changed for all tissues when the *APOE* locus was removed.

### PRS calculation

Each SNP set as defined above was then used to construct the corresponding tissue-specific PRS. Each PRS was constructed with PRSice[41] (v 2.2.4) using the Kunkle et al. 2019 IGAP summary statistics[7] as the base data set and the 1000 Genomes CEU samples to estimate linkage disequilibrium (LD). An R^2^ of 0.5 was used for clumping SNPs and a P threshold of 0.0025 based on the Zhao et al. 2019 estimation of the most predictive threshold for AD[42] was used for the inclusion of SNPs, leaving a maximum of 649,987 SNPs remaining for PRS construction depending on the SNP set used. The risk score for each tissue was calculated for each participant in the combined WADRC/WRAP data set using the default “average” PRS equation that divides the weighted sum of the alleles by the total count of alleles used (“--score avg” option for PRSice), and then each tissue-specific PRS was standardized to a mean of 0 and variance of 1.

### PRS-AD diagnosis associations

Among the WADRC/WRAP data set, all 1,410 participants had at least one study visit with a consensus conference diagnosis. The most recent visit for each participant with an AD, MCI, or cognitively unimpaired diagnosis was kept. Then, among related individuals (defined by estimated genetic relationships in WADRC or self-defined families in WRAP) only one participant was kept per family (chosen arbitrarily), leaving 1,164 unrelated participants. The association between each PRS and AD diagnosis (compared to cognitively unimpaired individuals; n = 79 cases with AD and n = 1,037 cognitively unimpaired controls; MCI cases excluded) was estimated with logistic regression using the R[43] glm function, controlling for sex and age at the time of diagnostic assessment by the consensus review committee. A Bonferroni correction for the number of PRS tested (P = 0.05 / 14 = 0.0036) was used in reporting significant results.

As a follow-up analysis, a comparison of the top-performing risk score between AD, MCI, and cognitively unimpaired participants was made using a box plot, with both ANOVA and pairwise t-tests used to compare the risk score distributions among the groups. Participants were then divided into 5 groups based on the PRS quantiles, and the distributions of the AD diagnoses across these PRS quantiles were compared.

Though the two cohort populations were similar geographically and demographically, the possibility of confounding by cohort or population substructure was assessed. The PRS-diagnosis associations were repeated as above but using only the participants from WADRC and including the first 5 principal components (PCs) of genetics derived previously using PC-AiR[44]. WADRC was used for this sensitivity analysis because it had a more balanced distribution of clinical diagnoses than WRAP.

To assess whether the effect of *APOE* was solely driving the associations of the PRS with AD diagnosis, a sensitivity analysis was conducted with the set of tissue-specific PRS constructed without the *APOE* locus. The associations between these PRS without *APOE* and AD diagnosis were estimated with logistic regression as before and compared to the original models with *APOE* included in the PRS.

To assess whether the top-performing PRS was substantially better than the rest of the genome in predicting AD diagnosis, a sensitivity analysis was conducted where only non-tissue-functional SNPs were used to construct a PRS. To build this PRS (referred to as an inverse PRS), the same procedure as before was used for the top-performing tissue PRS except that only SNPs in the non-functional regions for that tissue were included. The association of this PRS with AD diagnosis was estimated in a logistic regression as before and compared to that of the tissue-functional PRS, both with and without the *APOE* region included.

### PRS-CSF biomarker associations

To investigate the intermediate biological pathways driving the association between the PRS most strongly associated with AD diagnosis, the relationship between this PRS (both with and without *APOE*) and the CSF biomarkers was assessed using the longitudinal data set of unrelated WADRC and WRAP participants with biomarker measurements available (up to 250 visits from 167 individuals). A linear mixed-effects model was used to test each liver PRS-biomarker association, controlling for age at CSF collection and sex and including a random intercept for the individual to account for the longitudinal CSF measurements. A Bonferroni correction for the number of biomarkers tested (P = 0.05 / 9 = 0.0056) was used for reporting significant associations.

## Results

### PRS creation

A total of 14 PRS were initially generated based on the GenoSkyline-PLUS functionality annotation: one genome-wide PRS (labeled “all”) and 13 tissue-specific PRS (blood, thymus, and spleen; brain; breast; connective tissue; fat; gastrointestinal; heart; liver; lung; muscle; ovary; pancreas; and skin). The proportion of the 5,631,405 SNPs considered functional for each tissue ranged from 2.5% (ovary) to 25.3% (blood, thymus, and spleen) (Supplementary Figure 1). Among the 1,164 unrelated participants, the genome-wide and tissue-specific PRS values for the WADRC/WRAP cohort were all roughly normally distributed whether the *APOE* region was included or excluded from the PRS (Supplementary Figures 2-3). The PRS were generally correlated with each other with a median pairwise correlation of 0.872 (*APOE* included) and 0.754 (*APOE* excluded) (Supplementary Figures 4-5 and Supplementary Table 2).

### PRS-AD diagnosis association

The 1,164 unrelated participants were considered for the PRS-AD analyses (Table 1). The majority (1,037, 89.1%) of these participants were cognitively unimpaired at their most recent visit, with the AD and MCI diagnosis participants older and less often female. For the analysis of the PRS-AD diagnosis associations, only the 1,116 AD and cognitively unimpaired participants were used.

**Table 1.**
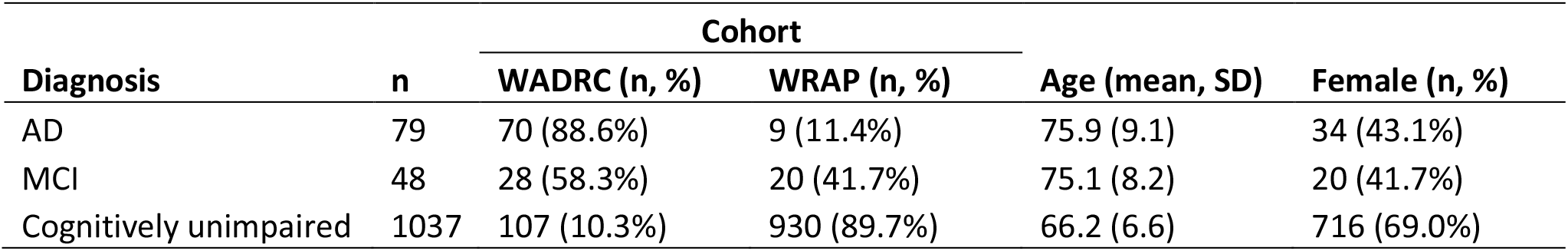
AD diagnosis cohort demographics.

The strength of association for each PRS with AD diagnosis (versus cognitively unimpaired) is shown in Figure 1 (full regression results and model areas under the curve [AUC] in Supplementary Table 3). All PRS were individually and statistically significantly associated with AD diagnosis after Bonferroni correction for multiple testing, with all PRS showing an increase of polygenic risk associated with a diagnosis of AD relative to cognitively unimpaired and AUCs between 0.839 and 0.866. Two tissues (liver and ovary) were more strongly associated with AD diagnosis than the genome-wide PRS, with an odds ratio of having AD relative to being cognitively impaired (OR) (and 95% confidence intervals [CI] and P values) of 2.19 per standard deviation (SD) increase in the PRS (95% CI = 1.70-2.82, P = 1.46 × 10^−9^) and 2.06 (1.59-2.66, P = 3.55 × 10^−8^), respectively, compared to the genome-wide PRS with OR of 2.01 (1.54-2.62, P = 2.40 × 10^−7^).

**Figure 1.**
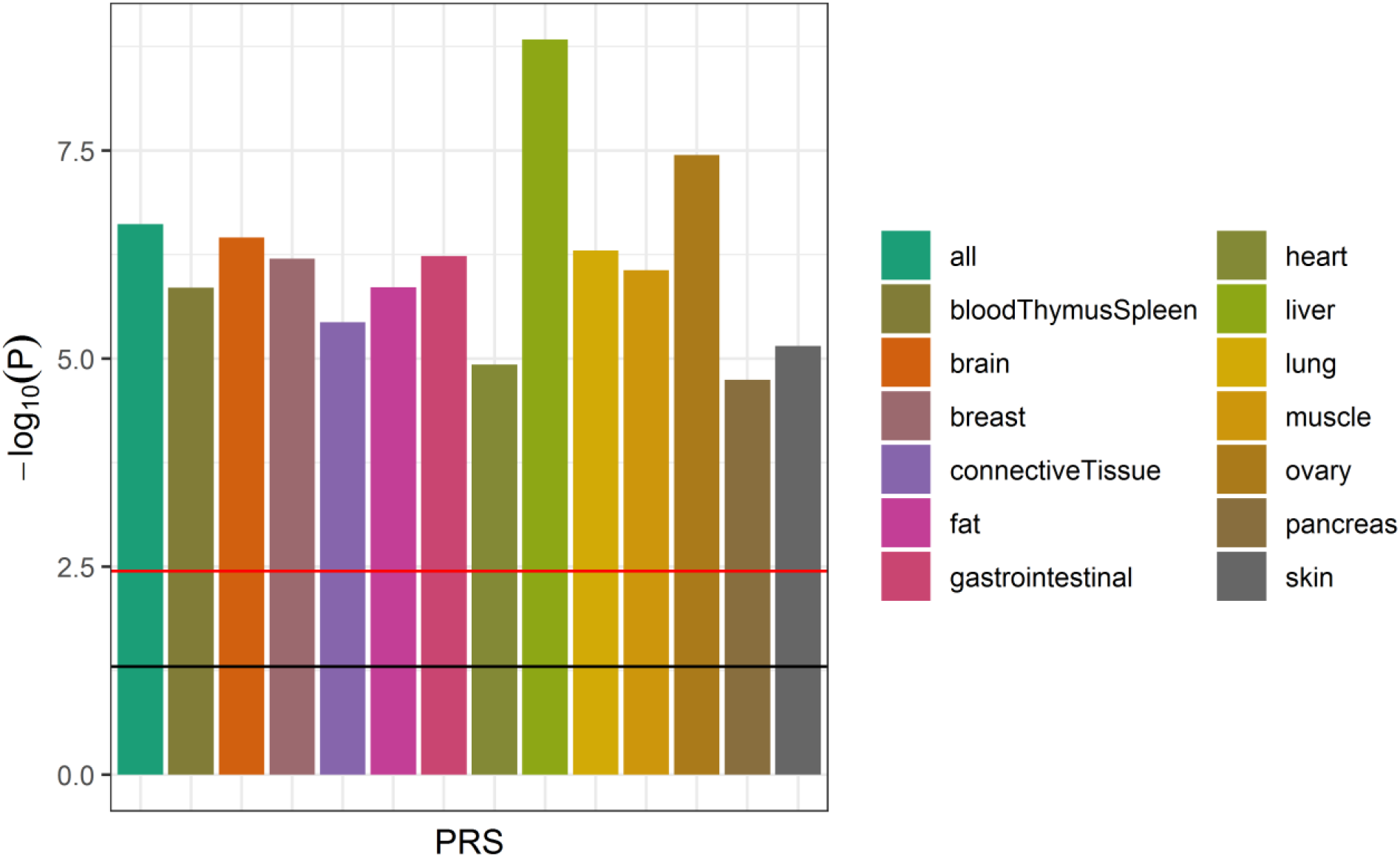
Association of tissue-specific PRS with AD diagnosis (*APOE* included) The strength of association of each PRS (with the *APOE* locus included) with AD diagnosis (relative to cognitively unimpaired participants; MCI cases excluded) from the logistic regression models is shown (n = 1,116). The horizontal lines indicate thresholds for significance, with the black line indicating the nominal threshold of P = 0.05 and the red line indicating the Bonferroni-corrected threshold of P = 0.0036. All PRS scores were statistically significantly associated with AD diagnosis, with the liver PRS being the most strongly associated score.

The genetic risk scores for the liver PRS (with *APOE*) increased with increasing severity across the three cognitive diagnoses from a mean value of -0.02 (SD = 0.96) for cognitively unimpaired individuals to 0.60 (SD = 1.10) for participants with AD (Figure 2). The mean liver score was significantly different across the three groups (ANOVA P = 1.6 × 10^−7^) and between the unimpaired and AD participants (t-test P = 4.8 × 10^−6^). When the participants were stratified into 5 liver PRS risk quantiles, the highest risk (5^th^ quantile) and lowest risk (1^st^ quantile) showed a corresponding enrichment of AD and MCI participants at the highest PRS risk (19.4%) relative to the lowest PRS risk (6.4%) (Supplementary Figure 6). Similar but less marked and consistent trends were seen for the liver PRS without *APOE*: the mean PRS value ranged from -0.001 (SD = 0.97) among cognitively unimpaired individuals to 0.34 (SD = 0.99) among participants with AD, and the proportion of AD and MCI participants among the highest risk quantile was 14.7% compared to 6.9% in the lowest risk quantile.

**Figure 2.**
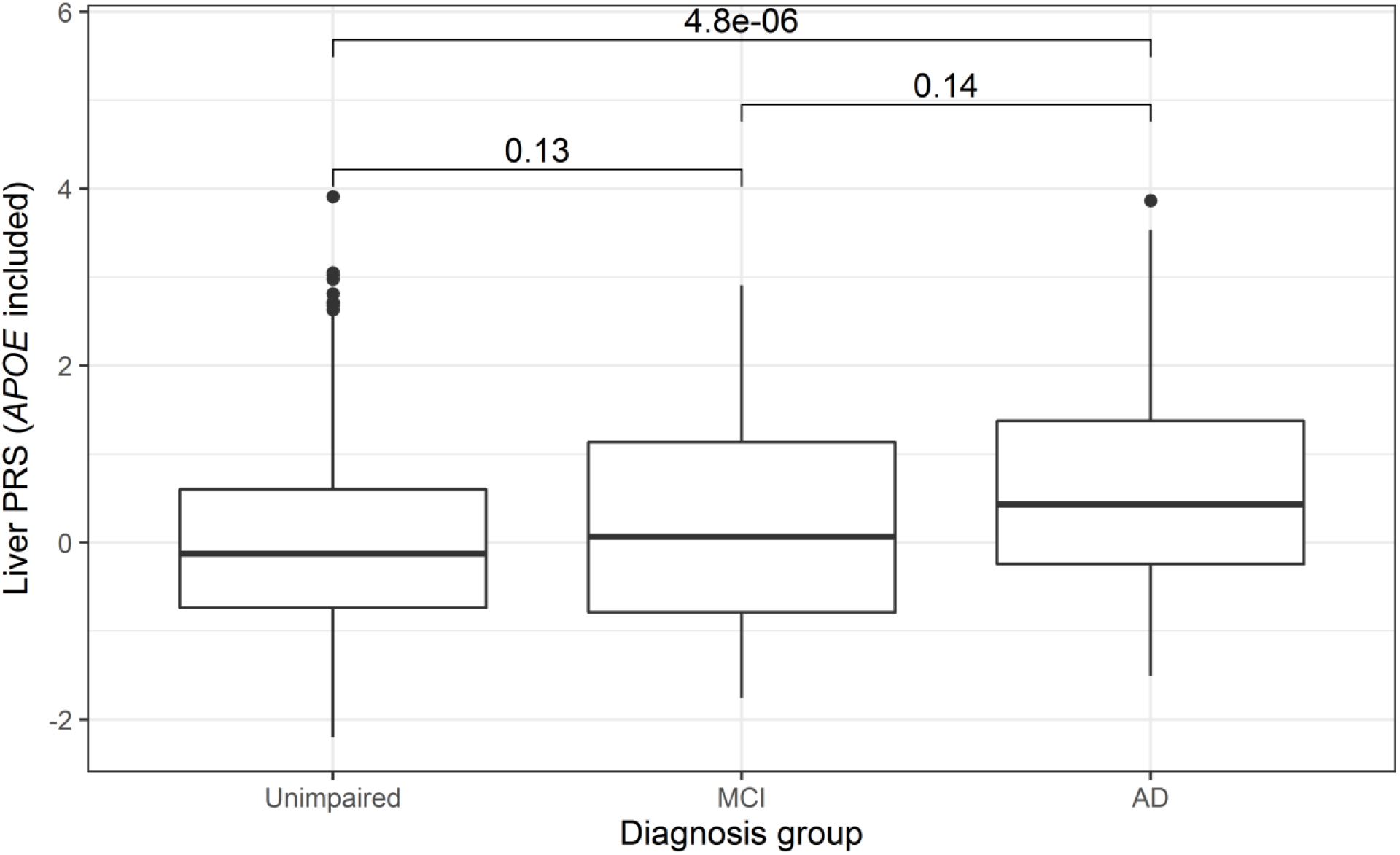
Distribution of liver PRS by diagnosis group (*APOE* included) The distribution of the liver PRS with the *APOE* region included across the three clinical stages of AD is shown (n = 1,410). Pairwise t-tests of the diagnosis group means (P values shown above each box plot pair) revealed a statistically significant difference between the liver PRS scores of the cognitively unimpaired and AD groups. An ANOVA test similarly identified a statistically significant difference in means across all three groups (P = 1.6 × 10^−7^).

When these PRS-AD diagnosis analyses were repeated using just the WADRC cohort (n = 177) and including the first 5 genetic PCs as additional covariates, the results were similar but with weaker associations. All PRS with *APOE* included were statistically significantly associated with AD diagnosis with the liver PRS being the most strongly associated with an OR of 2.26 (95% CI: 1.53-3.33, P = 3.86 × 10^−5^). When *APOE* was excluded, only the liver PRS remained statistically significantly associated with an OR of 1.95 (1.24-3.05, P = 0.00357), though its P value was just below the Bonferroni-corrected significance threshold.

To rule out the possibility that the PRS associations with AD diagnosis were solely driven by the effect of the *APOE* locus, the PRS-AD diagnosis associations were recalculated using the PRS that excluded the *APOE* region. In this sensitivity analysis, only the liver PRS remained significantly associated with AD diagnosis after Bonferroni correction for the number of PRS tested with an OR of 1.55 (95% CI: 1.19-2.02, P = 0.0012) (Figure 3), although the AUCs remained similar across PRS models (range: 0.811-0.823; Supplementary Table 3). A similar increasing genetic risk score across the diagnosis groups from cognitively unimpaired to AD was seen with the liver PRS without the *APOE* locus. A statistically significant difference in the genetic risks scores for the liver PRS between cognitively unimpaired (mean score = -0.00050, SD = 0.97) and AD (mean score = 0.34, SD = 0.99) groups was observed with a t-test P = 0.0041.

**Figure 3.**
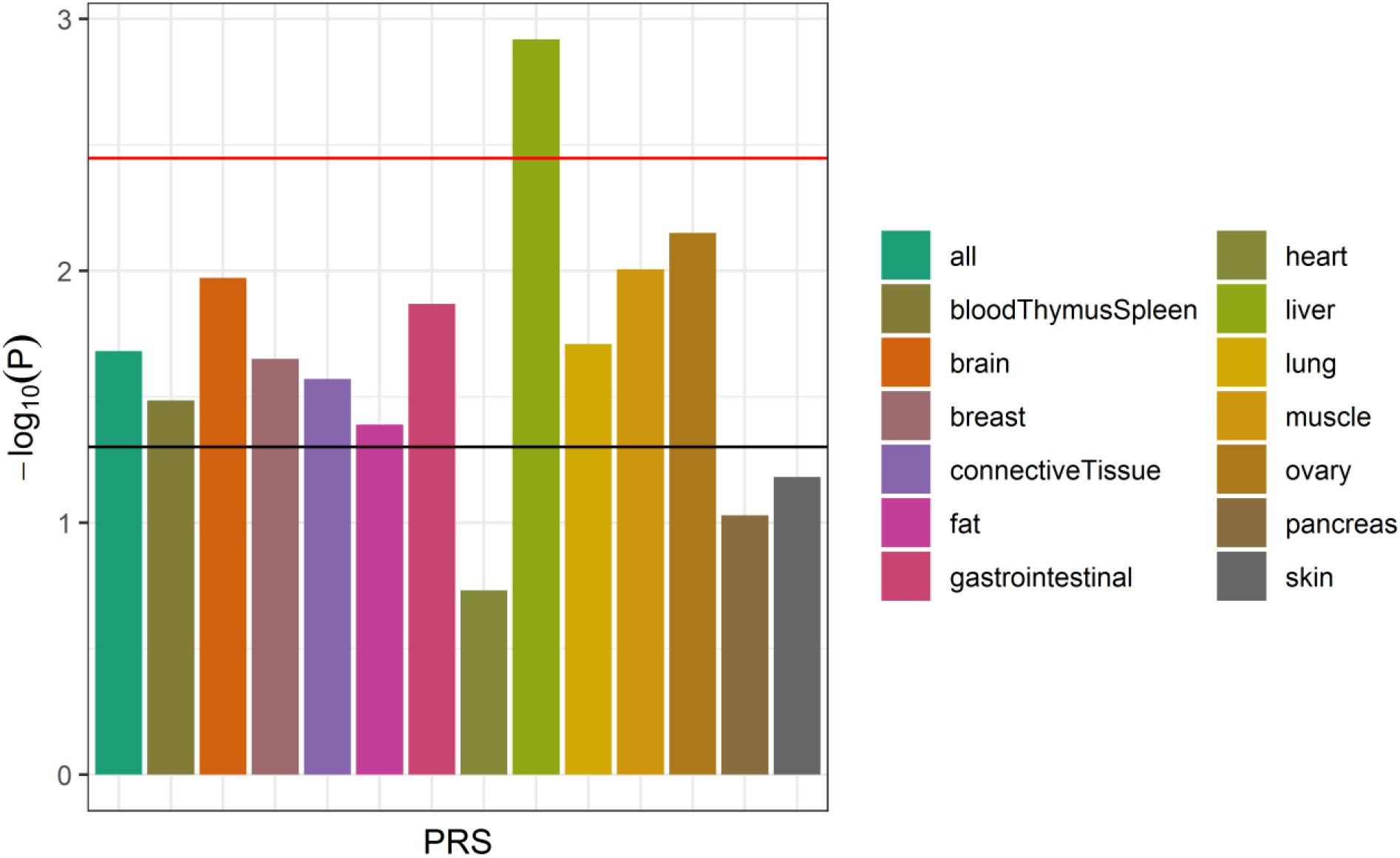
Association of tissue-specific PRS with AD diagnosis (*APOE* excluded) The strength of association of each PRS (with the *APOE* locus excluded) with AD diagnosis (relative to cognitively unimpaired participants; MCI cases excluded) from the logistic regression models is shown (n = 1,116). The horizontal lines indicate thresholds for significance, with the black line indicating the nominal threshold of P = 0.05 and the red line indicating the Bonferroni-corrected threshold of P = 0.0036. Only the liver PRS remained statistically significantly associated with AD diagnosis after multiple testing correction.

To explore whether the liver-functional genome was indeed more predictive of AD diagnosis than the remaining genome, the performance of the liver PRS was compared to the PRS constructed using every part of the genome except for the liver-functional genome (i.e., the “liver inverse” PRS), both with and without the *APOE* region included. Regardless of the inclusion of the *APOE* region, the liver-functional genome PRS was more strongly associated with AD diagnosis (vs. cognitively unimpaired) than the liver inverse PRS, although all PRS were nominally associated. With *APOE* included, the liver PRS’ OR was 2.19 (95% CI = 1.70-2.82, P = 1.46 × 10^−9^) compared to the liver inverse PRS with an OR of 1.88 (95% CI = 1.44-2.44, P = 2.92 × 10^−6^). Without *APOE*, the liver PRS OR was 1.55 (95% CI = 1.19-2.02, P = 0.0012) compared to the liver inverse PRS with an OR of 1.34 (95% CI = 1.04-1.72, P = 0.024) (Supplementary Figure 7).

### PRS-CSF biomarker associations

The longitudinal data set for the CSF biomarkers included all available WADRC/WRAP visits for participants where CSF biomarker and genetic data were available, which ranged from 1 to 5 visits per participant. The total number of participants included per biomarker analysis ranged from 164-167, comprising 245-250 total visits (Table 2). The mean age at visit across all included visits was 64.1 (SD 7.1) with 64.0% of the visits from female participants.

**Table 2.** CSF biomarker data set description.

| **Outcome** | **Individuals** | **Visits** | **Outcome mean** | **Outcome SD** |
| --- | --- | --- | --- | --- |
| α-synuclein (pg/mL) | 167 | 250 | 183.87 | 83.43 |
| Aβ42:Aβ40 ratio | 166 | 247 | 0.060 | 0.020 |
| IL-6 (pg/mL) | 164 | 245 | 4.48 | 2.62 |
| Neurogranin (pg/mL) | 167 | 250 | 849.62 | 354.44 |
| NFL (pg/mL) | 167 | 250 | 115.78 | 84.49 |
| ptau (pg/mL) | 166 | 247 | 21.59 | 12.55 |
| ptau:Aβ42 ratio | 166 | 247 | 0.034 | 0.034 |
| sTREM2 (ng/mL) | 167 | 250 | 8.74 | 2.82 |
| YKL-40 (ng/mL) | 167 | 250 | 160.05 | 63.76 |

The results of the linear mixed-effects models that regressed each outcome on the liver PRS (controlling for age at visit and sex) are summarized in Figure 4 (full regression results in Supplementary Table 4). After Bonferroni correction, the liver PRS was statistically significantly associated with three outcomes: the Aβ42/Aβ40 ratio, ptau, and the ptau/Aβ42 ratio. The liver PRS was nominally associated with three other outcomes: NFL, neurogranin, and α-synuclein. The remaining three outcomes (YKL-40, sTREM2, and IL-6) were not associated with the liver PRS.

**Figure 4.**
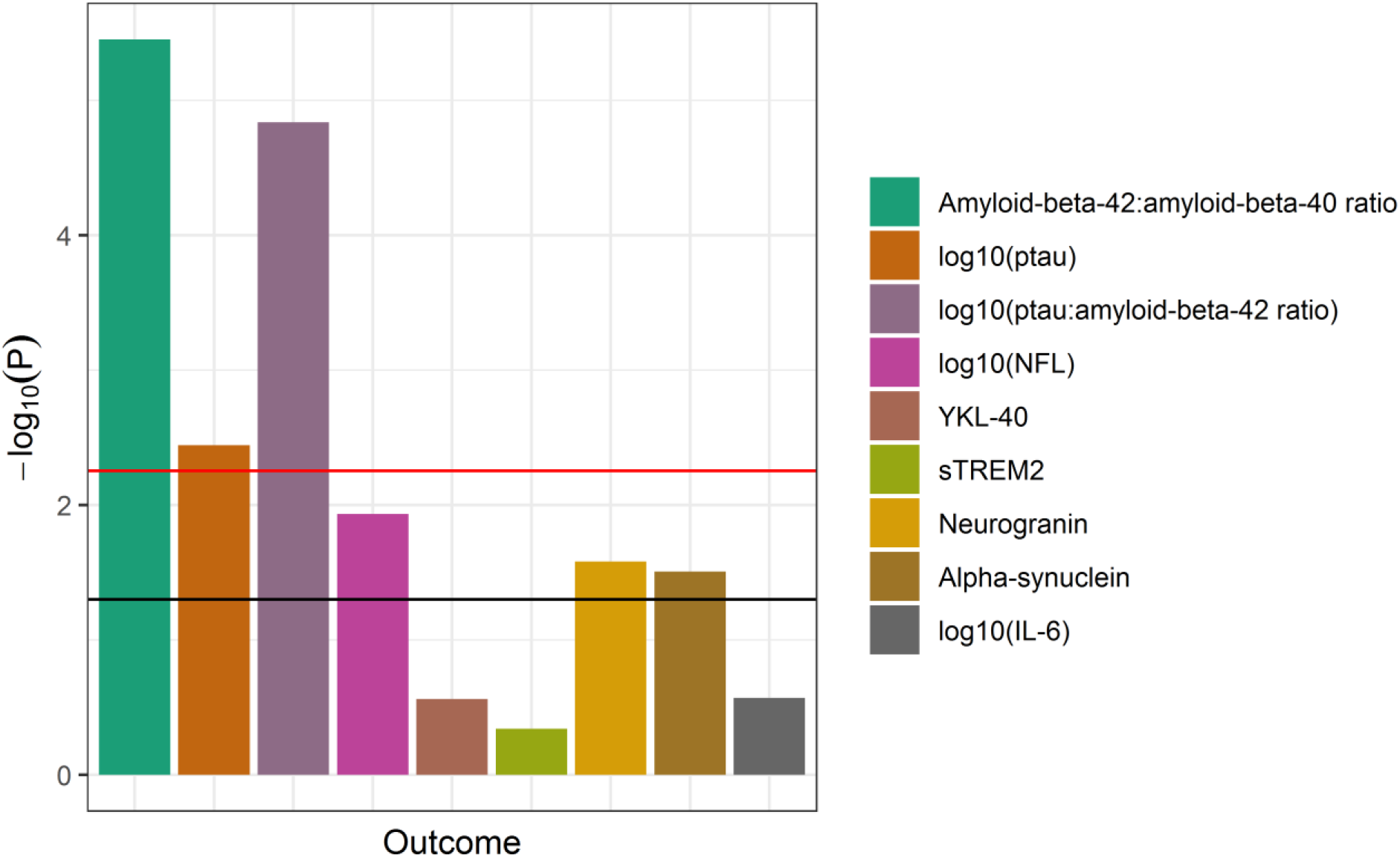
Association of the liver PRS with CSF biomarkers (*APOE* included) The strength of association of the liver PRS (with the *APOE* locus included) with each CSF biomarker from the linear mixed effects regression models is shown (n range = 245-250 visits). The horizontal lines indicate thresholds for significance, with the black line indicating the nominal threshold of P = 0.05 and the red line indicating the Bonferroni-corrected threshold of P = 0.0056. The liver PRS was statistically significantly associated with the measures of amyloid and tau but not with the other biomarkers.

When these analyses were repeated with the *APOE* region removed from the liver PRS, the majority of the association signal was lost: no outcome was statistically significantly associated with the liver PRS without *APOE* after Bonferroni correction, although the PRS was nominally associated with sTREM2 (Supplementary Figure 8).

## Discussion

The main result from the analysis of the association between tissue-specific PRS and AD diagnosis was that the liver PRS outperformed all other tissue-specific and non-tissue-specific PRS according to the strength of statistical association and the area under the curve (AUC), although the differences in AUC were subtle. When the effect of *APOE* was mitigated by excluding all SNPs in the *APOE* region, the liver PRS was the only PRS to remain statistically significant following multiple testing correction in its association to AD diagnosis. The importance of the *APOE* region in driving much of the association signal for the PRS models, including the liver PRS, was expected, as the *APOE* locus has long been known to be strongly associated with AD risk[45,46], especially among a population predominantly of European ancestry[47], as was the case here. However, the liver PRS was associated with AD diagnosis beyond the impact of the *APOE* locus.

Interpreting the meaning of the liver PRS’ relationship with AD was aided by the follow-up analysis with the CSF biomarkers. The liver PRS was most strongly associated with the core biomarkers of AD, amyloid and tau (CSF Aβ42/Aβ40, ptau, and ptau/Aβ42), suggesting that the PRS was more directly capturing these features of AD pathology rather than some of the other processes of neuroinflammation and neurodegeneration. However, these associations with amyloid and tau were removed when the *APOE* locus was removed from the liver PRS, leaving only a nominal association with sTREM2. Weaker association signals among the CSF biomarker data set could be attributed in part to the much smaller sample size available with CSF biomarker data compared to that with AD diagnosis data (n = 250 vs n = 1,116). Still, the reason for the liver PRS without *APOE* being associated with AD diagnosis but not any of the CSF biomarkers was unclear.

Whether the liver PRS’ association with AD risk indicates a role for the liver organ itself remains an open question. The liver PRS here may be associated with AD due to some role of the liver itself or simply through the genes that happen to be functional in the liver but are not uniquely expressed in the liver. Across the 560 SNPs that were part of the liver PRS (*APOE* excluded), many of the major AD loci were represented, including *CLU, BIN1, PICALM, SPI1, CLU, ABCA7, SORL1*, and others (Supplementary Table 5). Nevertheless, there is mounting evidence pointing to metabolic dysregulation and the liver as relevant to AD. Several metabolic traits, including dyslipidemia, metabolic syndrome, obesity, and type 2 diabetes, appear to be risk factors for AD[48,49]. More specific to the liver, Neuner et al. suggest that cholesterol regulation may be a point of common ground between the liver, *APOE*, neurons, and AD[6]. Furthermore, recent evidence has indicated an association between measures of liver function, including blood levels of alanine aminotransferase (ALT) and aspartate aminotransferase (AST), and AD diagnosis, amyloid, tau, and neurodegeneration[4]. Though the causal direction of these associations is unclear, our findings here provide potential further evidence of the relevance of the liver and metabolism in relation to AD.

More generally, this study reinforces the benefit of using functional annotation to improve genomic prediction as the PRS’ performance was improved, albeit subtly, by incorporating predicted functional information. Among the PRS models with *APOE* excluded, 6 of the 13 tissue-specific PRS were more strongly associated with AD diagnosis than the genome-wide PRS in association. This increased strength of association is likely the result of improved filtering of the included SNPs to just those that are more likely to be causal due to their predicted functionality. This finding would support a general theme among the functional annotation literature that suggests that functional annotation can improve genomic analyses of disease. Early work demonstrated that genomic functional annotation could be used to filter down a set of SNPs to those more likely to be causal for both dominant and recessive Mendelian traits[50]. Recent approaches have used functional annotation to boost GWAS power in identifying SNP associations[51], stratify heritability of complex disease by functional annotation[52], and improve genetic risk prediction for disease[20,53]. Our work further demonstrates the potential utility of incorporating functional annotation in genetic risk prediction, though additional work is needed to quantify whether the improvement in this case is enough to be clinically relevant.

Limitations of this study included the limited sample size. In the study of PRS-AD diagnosis associations, the sample was predominantly cognitively unimpaired with only 79 individuals diagnosed with AD, and in the follow-up analysis of CSF biomarker data, only 167 unique individuals were available with data. However, even among these smaller sample sizes, detectable association signals were still observed. As these cohort studies continue to grow, so too will our capability to investigate genetic associations with AD pathology. Another limitation was the population of study, which was limited to European ancestry due to lack of participants from other populations in the data set at the time the data were pulled. Further studies will be needed to better understand the transferability of these tissue-specific PRS findings to other populations.

In conclusion, we leveraged genome functional annotation to create tissue-specific PRS for AD, identifying the liver PRS as the PRS most strongly associated with AD and the only PRS to remain associated when the *APOE* locus was removed. Follow-up analysis of the liver PRS with CSF biomarkers of AD, neurodegeneration, and neuroinflammation revealed potential intermediate pathways related to the role of the liver-functional genome in AD, but the limited sample size of the biomarker data set and the apparent role of *APOE* in driving these biomarker results merit further study. Altogether, these findings provide further evidence for the role of the liver-functional genome in AD and highlight the benefit of incorporating genomic functional annotation into genetic research of complex disease.

## Supporting information

STREGA checklist

## Data Availability

The datasets analyzed in this study may be requested from the WADRC at https://www.adrc.wisc.edu/apply-resources.

## Acknowledgments

We would like to thank WRAP and WADRC participants and the Wisconsin Alzheimer’s Institute (WAI) and WADRC staff for their contributions to the WRAP and WADRC studies. Without their efforts this research would not be possible. This research is supported by National Institutes of Health (NIH) grants R01AG27161 (Wisconsin Registry for Alzheimer Prevention: Biomarkers of Preclinical AD), R01AG054047 (Genomic and Metabolomic Data Integration in a Longitudinal Cohort at Risk for Alzheimer’s Disease), R21AG067092 (Identifying Metabolomic Risk Factors in Plasma and Cerebrospinal Fluid for Alzheimer’s Disease), R01AG037639 (White Matter Degeneration: Biomarkers in Preclinical Alzheimer’s Disease), P30AG017266 (Center for Demography of Health and Aging), and P50AG033514 and P30AG062715 (Wisconsin Alzheimer’s Disease Research Center Grant), the Helen Bader Foundation, Northwestern Mutual Foundation, Extendicare Foundation, State of Wisconsin, the Clinical and Translational Science Award (CTSA) program through the NIH National Center for Advancing Translational Sciences (NCATS) grant UL1TR000427, and the University of Wisconsin-Madison Office of the Vice Chancellor for Research and Graduate Education with funding from the Wisconsin Alumni Research Foundation. This research was supported in part by the Intramural Research Program of the National Institute on Aging. Computational resources were supported by a core grant to the Center for Demography and Ecology at the University of Wisconsin-Madison (P2CHD047873). We also acknowledge use of the facilities of the Center for Demography of Health and Aging at the University of Wisconsin-Madison, funded by NIA Center grant P30AG017266. Author DJP was supported by an NLM training grant to the Bio-Data Science Training Program (T32LM012413). Author BFD was supported by an NLM training grant to the Computation and Informatics in Biology and Medicine Training Program (NLM 5T15LM007359). Author YKD was supported by a training grant from the National Institute on Aging (T32AG000213). Author KB was supported by the Swedish Research Council (#2017-00915), the Alzheimer Drug Discovery Foundation (ADDF), USA (#RDAPB-201809-2016615), the Swedish Alzheimer Foundation (#AF-742881), Hjärnfonden, Sweden (#FO2017-0243), the Swedish state under the agreement between the Swedish government and the County Councils, the ALF-agreement (#ALFGBG-715986), the European Union Joint Program for Neurodegenerative Disorders (JPND2019-466-236), and the NIH, USA, (grant #1R01AG068398-01).

ELECSYS, COBAS and COBAS E are trademarks of Roche. The Roche NeuroToolKit robust prototype assays are for investigational purposes only and are not approved for clinical use.

We thank the University of Wisconsin Madison Biotechnology Center Gene Expression Center for providing Illumina Infinium genotyping services. We thank Dr. Brian Kunkle for his help with discussions on the sample overlap issue of genomic cohorts.

The content is solely the responsibility of the authors and does not necessarily represent the official views of the NIH.

## Data availability

The data sets analyzed in this study may be requested from the WADRC at https://www.adrc.wisc.edu/apply-resources.

