## Supplementary Figures for "Liver-specific polygenic risk score is more strongly associated than genome-wide score with Alzheimer’s disease diagnosis in a case-control analysis"

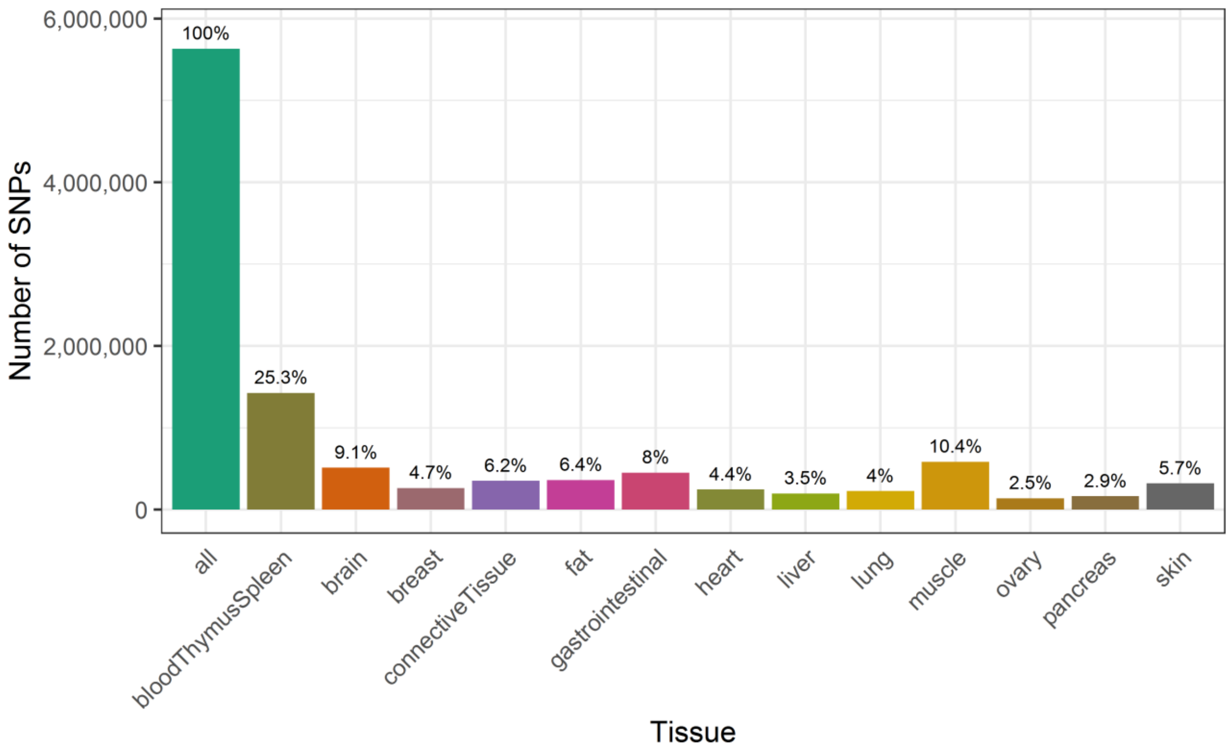

**Supplementary Figure 1. Functional SNPs by tissue**

The total number of SNPs from the IGAP GWAS summary statistics as well as the number of functional SNPs included for each tissue are shown. For most tissues, less than 10% of the total SNPs available from the IGAP GWAS were predicted to be functional for that tissue according to GenoSkyline-PLUS. The exception was the blood, thymus, and spleen tissue, which included more than a quarter of the total IGAP SNPs.

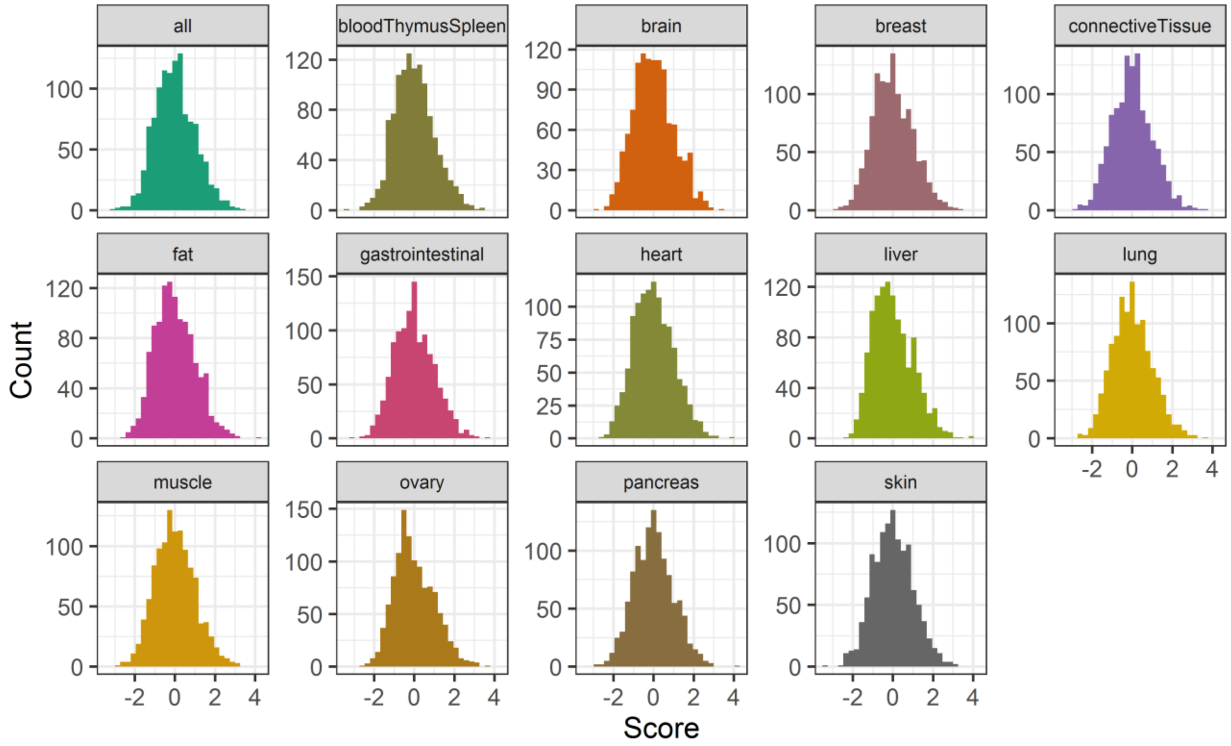

**Supplementary Figure 2. PRS score distribution by tissue (*APOE* included)**

The distribution of the calculated genetic risk scores for each tissue among the unrelated WADRC/WRAP data set (n = 1,164), with the *APOE* locus permitted to be included. Each tissue-specific score was roughly normally distributed.

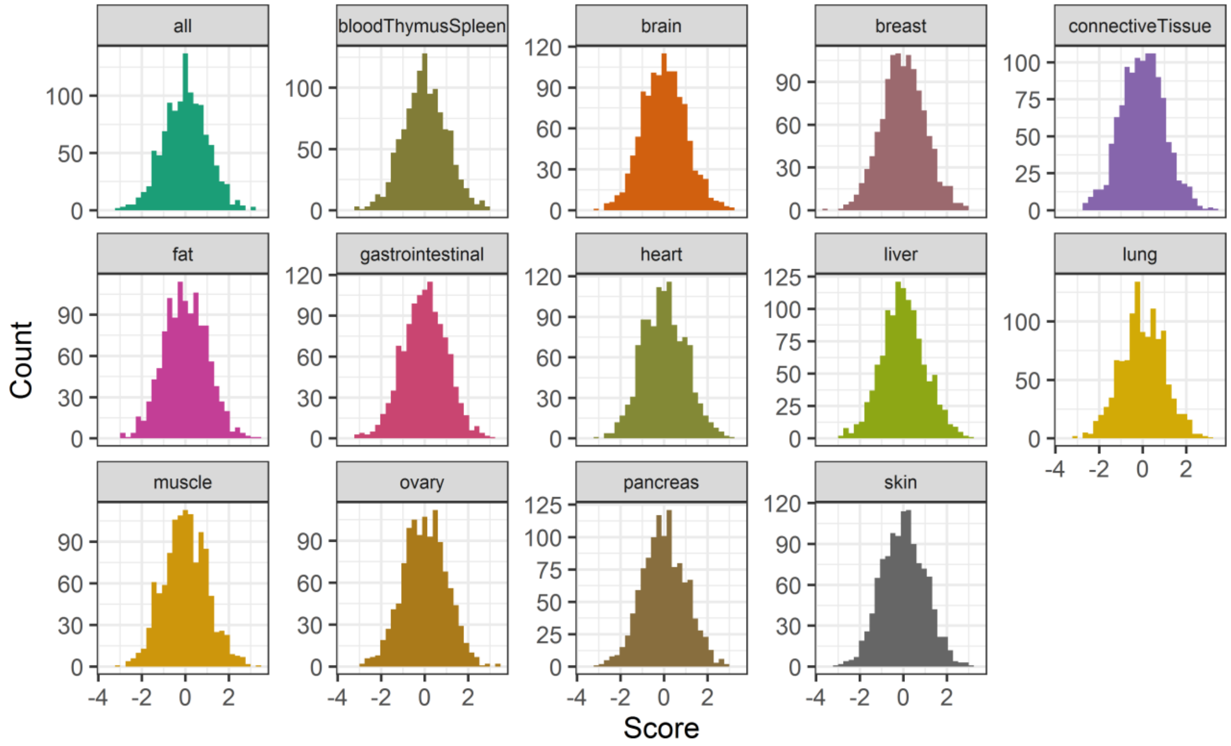

**Supplementary Figure 3. PRS score distribution by tissue (*APOE* excluded)**

The distribution of the calculated genetic risk scores for each tissue among the unrelated WADRC/WRAP data set (n = 1,164), with the *APOE* locus excluded. Each tissue-specific score was roughly normally distributed.

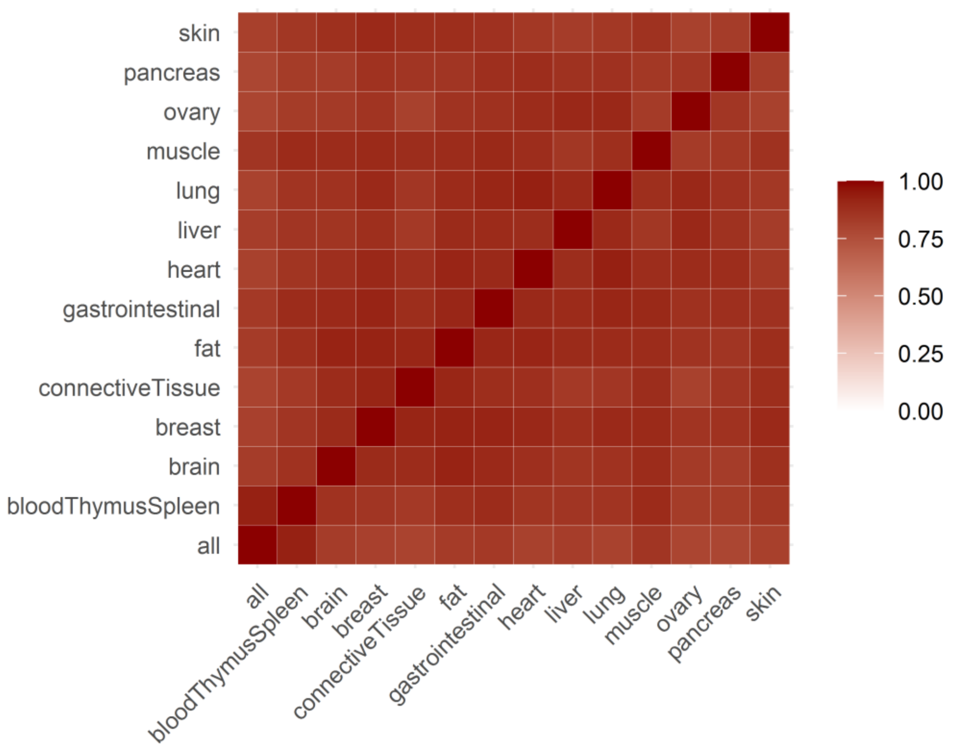

**Supplementary Figure 4. Correlation heat map of PRS scores (*APOE* included)**

The correlation of the calculated genetic risk scores between each tissue among the unrelated WADRC/WRAP data set (n = 1,164), with the *APOE* locus permitted to be included. All tissue PRS were strongly correlated with each other, which potentially reflected the relative importance of the *APOE* locus in driving PRS scores.

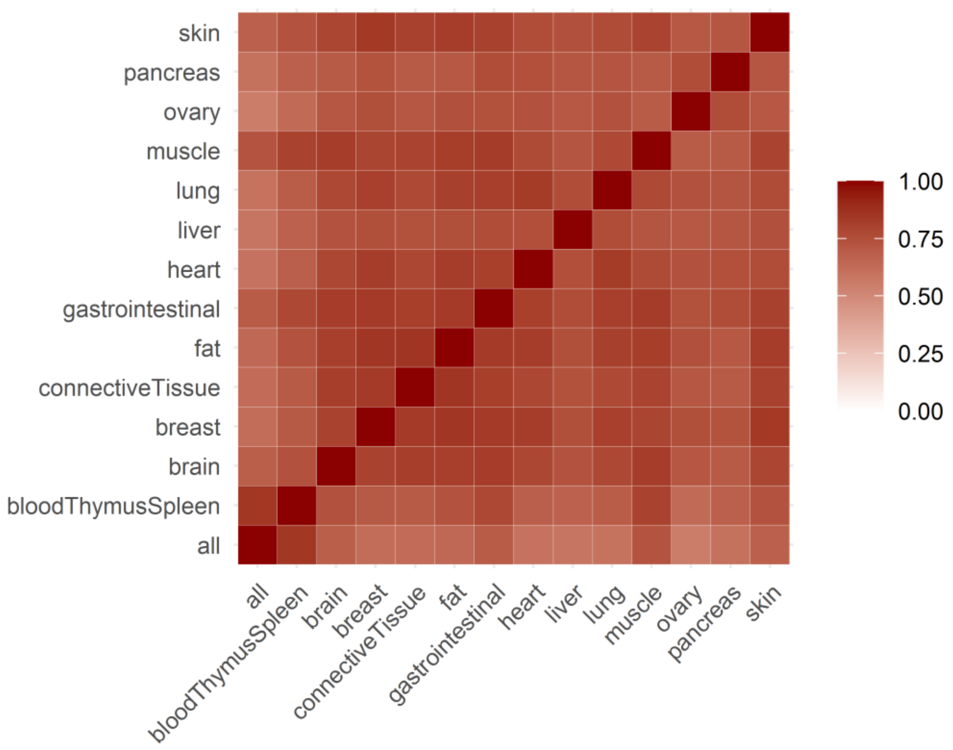

**Supplementary Figure 5. Correlation heat map of PRS scores (*APOE* excluded)**

The correlation of the calculated genetic risk scores between each tissue among the unrelated WADRC/WRAP data set (n = 1,164), with the *APOE* locus excluded. All tissue PRS remained strongly correlated with each other, although the correlations were lesser compared to those among the PRS with *APOE* included.

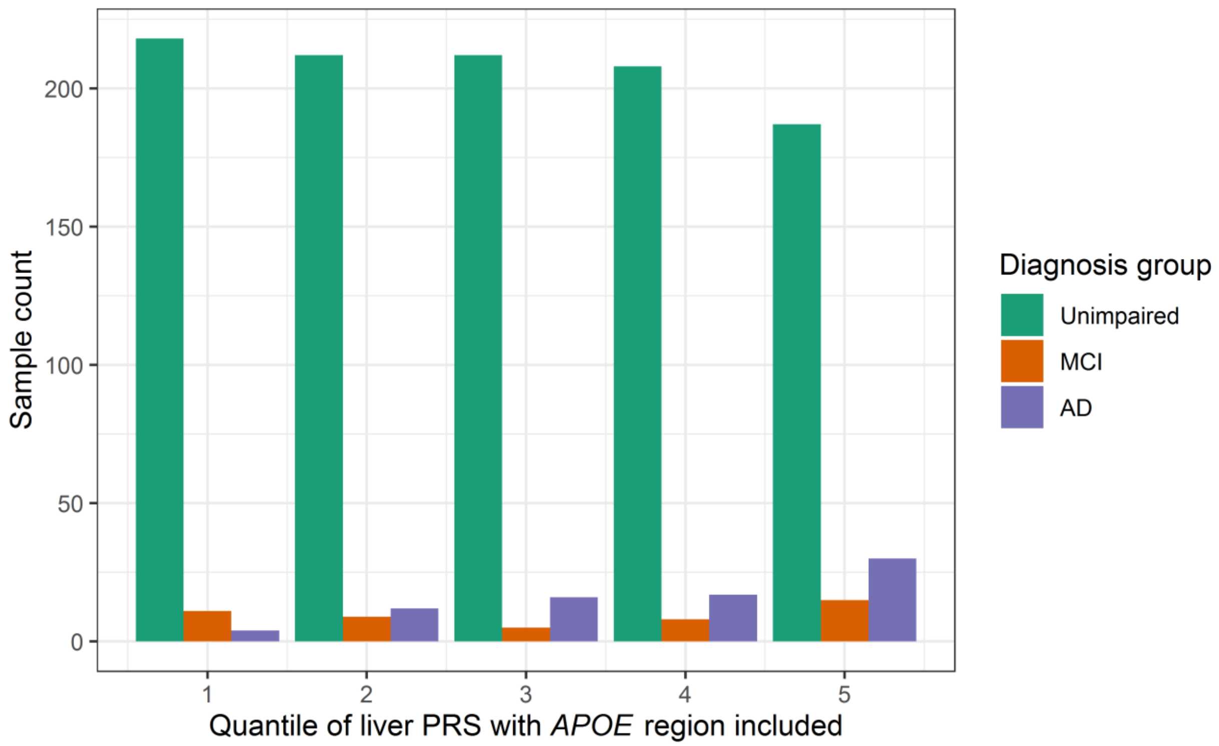

**Supplementary Figure 6. Distribution of diagnosis groups across liver PRS quantiles (*APOE* included)**

The distributions of the diagnosis groups across the quantiles (split into quintiles) of the liver PRS (with the *APOE* locus included) are shown. A decrease in the proportion of cognitively unimpaired individuals and an increase in the proportion of individuals diagnosed with AD can be seen with increasing genetic risk from the liver PRS.

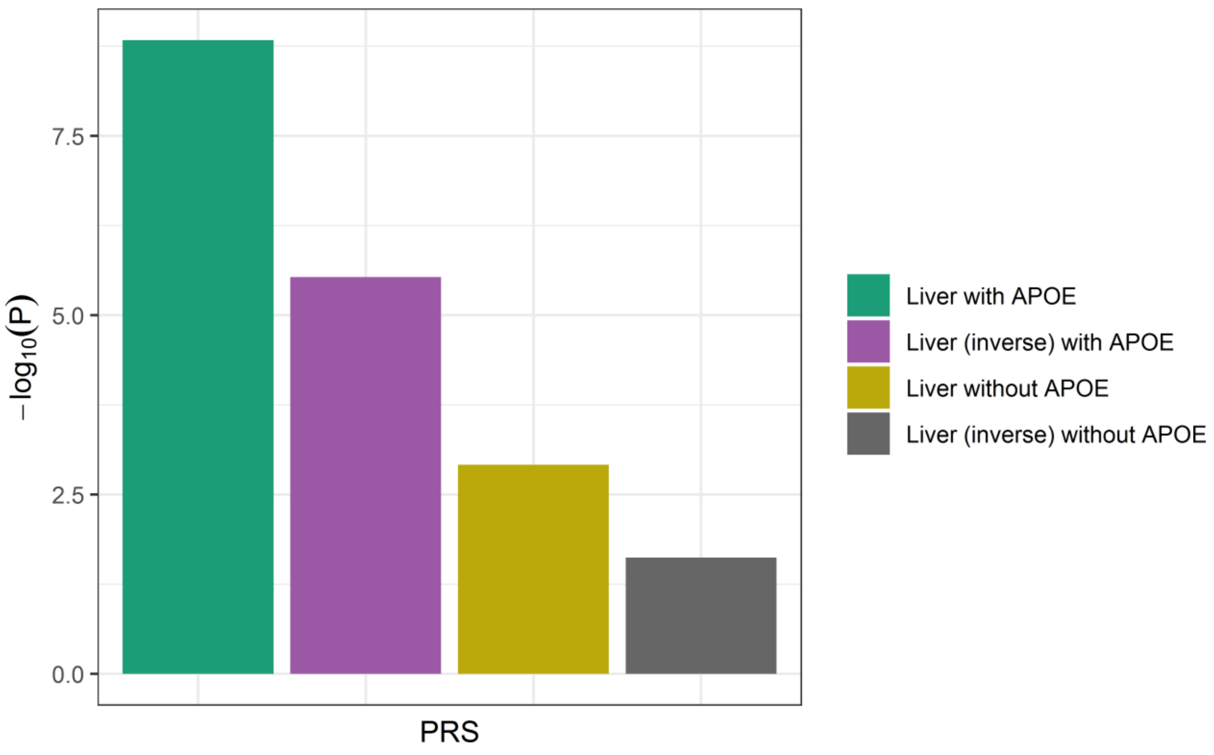

**Supplementary Figure 7. Relative strength of association of the liver PRS**

The strength of association between different variations of the liver PRS and AD diagnosis are summarized.

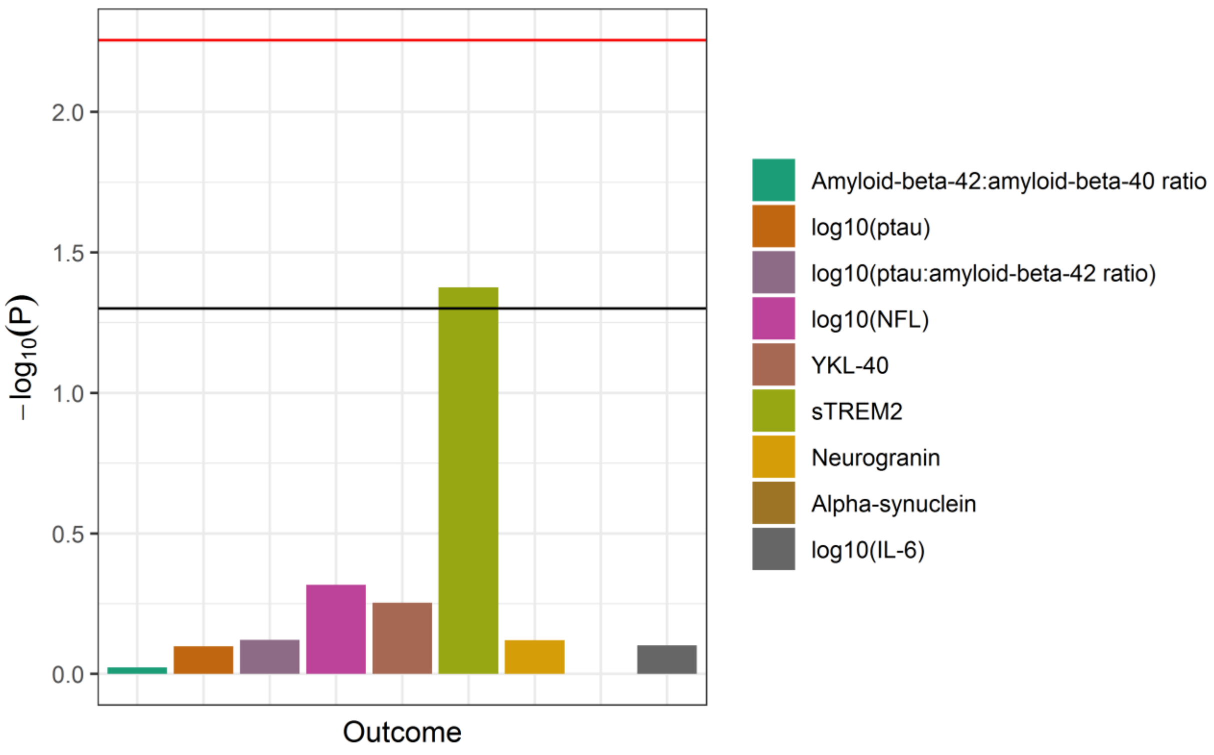

**Supplementary Figure 8. Association of the liver PRS with CSF biomarkers (*APOE* excluded)**

The strength of association of the liver PRS (with the *APOE* locus excluded) with each CSF biomarker from the linear mixed effects regression models is shown (n range = 245-250 visits). The horizontal lines indicate thresholds for significance, with the black line indicating the nominal threshold of P = 0.05 and the red line indicating the Bonferroni-corrected threshold of P = 0.0056. The liver PRS was only nominally associated with sTREM2.
