## Supplementary material for "Liver-specific polygenic risk score is more strongly associated than genome-wide score with Alzheimer’s disease diagnosis in a case-control analysis": STREGA checklist

### Reporting checklist for genetic association study.

Based on the STREGA guidelines.

#### Instructions to authors

Complete this checklist by entering the page numbers from your manuscript where readers will find each of the items listed below.

Your article may not currently address all the items on the checklist. Please modify your text to include the missing information. If you are certain that an item does not apply, please write "n/a" and provide a short explanation.

Upload your completed checklist as an extra file when you submit to a journal.

In your methods section, say that you used the STREGAreporting guidelines, and cite them as:

Little J, Higgins JP, Ioannidis JP, Moher D, Gagnon F, von Elm E, Khoury MJ, Cohen B, Davey-Smith G, Grimshaw J, Scheet P, Gwinn M, Williamson RE, Zou GY, Hutchings K, Johnson CY, Tait V, Wiens M, Golding J, van Duijn C, McLaughlin J, Paterson A, Wells G, Fortier I, Freedman M, Zecevic M, King R, Infante-Rivard C, Stewart A, Birkett N; STrengthening the REporting of Genetic Association Studies. STrengthening the REporting of Genetic Association Studies (STREGA): An Extension of the STROBE Statement.

| | | Reporting Item | Page Number |
|---|---|---|---|
| **Title and abstract** | | | |
| Title | #1a | Indicate the study's design with a commonly used term in the title or the abstract | p. 1 |
| Abstract | #1b | Provide in the abstract an informative and balanced summary of what was done and what was found | p. 2 |
| **Background/rationale** | | | |
| | #2 | Explain the scientific background and rationale for the investigation being reported | p. 3 |
| **Objectives** | | | |

| | | | |
|---|---|---|---|
| | #3 | State specific objectives, including any prespecified hypotheses. State if the study is the first report of a genetic association, a replication effort, or both. | p. 3. |
| **Study design** | | | |
| | #4 | Present key elements of study design early in the paper | p. 3. The end of the intro presents the key elements of the study design, which are further detailed in the methods (pp. 3-7). |
| **Setting** | | | |
| | #5 | Describe the setting, locations, and relevant dates, including periods of recruitment, exposure, follow-up, and data collection | pp. 3-4. A brief overview of the study cohorts is given, with citations included with further descriptions of the cohorts. |
| **Eligibility criteria** | | | |
| | #6a | Cohort study – Give the eligibility criteria, and the sources and methods of selection of participants. Describe methods of follow-up. Case-control study – Give the eligibility criteria, and the sources and methods of case ascertainment and control selection. Give the rationale for the choice of cases and controls. Cross-sectional study – Give the eligibility criteria, and the sources and methods of selection of participants. Give information on the criteria and methods for selection of subsets | pp. 3-7. A brief overview of the recruitment of participants and eligibility is provided, with citations to papers with additional information on the cohorts. Specific criteria used in this study to select the relevant subgroup of participants to analyze are provided throughout the Methods. |

| | | | |
|---|---|---|---|
| | | of participants from a larger study, when relevant. | |
| | #6b | Cohort study – For matched studies, give matching criteria and number of exposed and unexposed. Case-control study – For matched studies, give matching criteria and the number of controls per case. | p. 8. Table 1 provides a detailed breakdown of numbers. No matching was used. |
| **Variables** | | | |
| | #7a | Clearly define all outcomes, exposures, predictors, potential confounders, and effect modifiers. Give diagnostic criteria, if applicable | pp. 3-7. The diagnosis process is described with a citation to a more detailed description. All other outcomes (namely, CSF biomarkers), predictors, and confounders are described. |
| | #7b | Clearly define genetic exposures (genetic variants) using a widely-used nomenclature system. Identify variables likely to be associated with population stratification (confounding by ethnic origin). | The process of creating the PRS is described in detail in the Methods (pp. 5-6). When specific genes are referenced, gene names from GENCODE are used. Only a single population was used (European ancestry), which is described as a limitation in the results section (pp. 14-15). The potential impact of population and cohort is assessed as a sensitivity analysis (pp. 7, 10). |
| **Data sources/measurement** | | | |
| | #8a | For each variable of interest give sources of data and details of methods of assessment (measurement). Describe comparability of assessment | pp. 3-7 (Methods) |

| | | | |
|---|---|---|---|
| | | methods if there is more than one group. Give information separately for for exposed and unexposed groups if applicable. | |
| | #8b | Describe laboratory methods, including source and storage of DNA, genotyping methods and platforms (including the allele calling algorithm used, and its version), error rates and call rates. State the laboratory / centre where genotyping was done. Describe comparability of laboratory methods if there is more than one group. Specify whether genotypes were assigned using all of the data from the study simultaneously or in smaller batches. | pp. 3-7 (Methods) and provided citations. |
| **Bias** | | | |
| | #9a | Describe any efforts to address potential sources of bias | Age and sex are included as covariates. Cohort and population substructure are assessed in a sensitivity analysis (pp. 7, 10). |
| | #9b | Describe any efforts to address potential sources of bias | Age and sex are included as covariates. Cohort and population substructure are assessed in a sensitivity analysis (pp. 7, 10). |
| **Study size** | | | |
| | #10 | Explain how the study size was arrived at | pp. 3-7 (Methods) |
| **Quantitative variables** | | | |

| | | | |
|---|---|---|---|
| | #11 | Explain how quantitative variables were handled in the analyses. If applicable, describe which groupings were chosen, and why. If applicable, describe how effects of treatment were dealt with. | pp. 3-7 (Methods) |
| **Statistical methods** | | | |
| | #12a | Describe all statistical methods, including those used to control for confounding. State software version used and options (or settings) chosen. | pp. 3-7 (Methods) |
| | #12b | Describe any methods used to examine subgroups and interactions | N/A. No subgroup or interaction analyses were performed beyond the cohort-specific sensitivity analysis (p. 7). |
| | #12c | Explain how missing data were addressed | pp. 3-7 (Methods). Imputation was used for the genomic data. Otherwise, only non-missing data were used. |
| | #12d | If applicable, explain how loss to follow-up was addressed | N/A. This study does not address loss to follow-up. |
| | #12e | Describe any sensitivity analyses | pp. 3-7 (Methods) |
| | #12f | State whether Hardy-Weinberg equilibrium was considered and, if so, how. | p. 5 |
| | #12g | Describe any methods used for inferring genotypes or haplotypes | p.5 |

| | | | |
|---|---|---|---|
| | #12h | Describe any methods used to assess or address population stratification. | Cohort and population substructure are assessed in a sensitivity analysis (pp. 7, 10). |
| | #12i | Describe any methods used to address multiple comparisons or to control risk of false positive findings. | pp. 3-7 (Methods) |
| | #12j | Describe any methods used to address and correct for relatedness among subjects | pp. 3-7 (Methods) |
| **Participants** | | | |
| | #13a | Report numbers of individuals at each stage of study—eg numbers potentially eligible, examined for eligibility, confirmed eligible, included in the study, completing follow-up, and analysed. Give information separately for for exposed and unexposed groups if applicable. Report numbers of individuals in whom genotyping was attempted and numbers of individuals in whom genotyping was successful. | pp. 3-7 (Methods) |
| | #13b | Give reasons for non-participation at each stage | N/A. This study does not address reasons for non-participation in the underlying cohorts, though cited papers provide additional details on the cohorts. |
| | #13c | Consider use of a flow diagram | N/A. |
| **Descriptive data** | | | |
| | #14a | Give characteristics of study participants (eg demographic, | Key demographics are shown in Table 1 (p. 8). |

| | | | |
|---|---|---|---|
| | | clinical, social) and information on exposures and potential confounders. Give information separately for exposed and unexposed groups if applicable. Consider giving information by genotype | |
| | #14b | Indicate number of participants with missing data for each variable of interest | Discussion of missingness for genetic data is on p. 5. Missingness of other variables, such as individual CSF biomarkers, can be seen in the differences in sample size by each biomarker (Table 2, p.11). |
| | #14c | Cohort study – Summarize follow-up time, e.g. average and total amount. | N/A. Cohorts are ongoing. Further details are in the cited papers. |
| **Outcome data** | | | |
| | #15 | Cohort study Report numbers of outcome events or summary measures over time.Give information separately for exposed and unexposed groups if applicable. Report outcomes (phenotypes) for each genotype category over time Case-control study – Report numbers in each exposure category, or summary measures of exposure.Give information separately for cases and controls . Report numbers in each genotype category. Cross-sectional study – Report numbers of outcome events or summary measures. Give information separately for | p. 8 with Table 1 and p.11 with Table 2. Breakdown of diagnosis by PRS quantile in Sup. Fig. 6. |

| | | | |
|---|---|---|---|
| | | exposed and unexposed groups if applicable. Report outcomes (phenotypes) for each genotype category | |
| **Main results** | | | |
| | #16a | Give unadjusted estimates and, if applicable, confounder-adjusted estimates and their precision (eg, 95% confidence interval). Make clear which confounders were adjusted for and why they were included | Only appropriately confounder-adjusted estimates are provided. Included confounders are described (p. 6). |
| | #16b | Report category boundaries when continuous variables were categorized | p. 7 |
| | #16c | If relevant, consider translating estimates of relative risk into absolute risk for a meaningful time period | N/A. |
| | #16d | Report results of any adjustments for multiple comparisons | Reported throughout methods and results with Bonferroni corrections. |
| **Other analyses** | | | |
| | #17a | Report other analyses done—e.g., analyses of subgroups and interactions, and sensitivity analyses | Multiple sensitivity and subgroup analyses are reported throughout the paper. |
| | #17b | Report other analyses done—e.g., analyses of subgroups and interactions, and sensitivity analyses | Multiple sensitivity and subgroup analyses are reported throughout the paper. |
| | #17c | Report other analyses done—e.g., analyses of subgroups and interactions, and sensitivity analyses | N/A. All analyses relevant to this particular paper are either reported or cited or not yet published elsewhere. |

##### Key results

| | | | |
|---|---|---|---|
| | #18 | Summarise key results with reference to study objectives | pp. 7-13. |

##### Limitations

| | | | |
|---|---|---|---|
| | #19 | Discuss limitations of the study, taking into account sources of potential bias or imprecision. Discuss both direction and magnitude of any potential bias. | pp. 14-15 |

##### Interpretation

| | | | |
|---|---|---|---|
| | #20 | Give a cautious overall interpretation considering objectives, limitations, multiplicity of analyses, results from similar studies, and other relevant evidence. | pp. 13-15 |

##### Generalisability

| | | | |
|---|---|---|---|
| | #21 | Discuss the generalisability (external validity) of the study results | pp. 13-15 |

##### Funding

| | | | |
|---|---|---|---|
| | #22 | Give the source of funding and the role of the funders for the present study and, if applicable, for the original study on which the present article is based | pp. 15-16 |

None The STREGA checklist is distributed under the terms of the Creative Commons Attribution License CC-BY. This checklist can be completed online using https://www.goodreports.org/, a tool made by the EQUATOR Network in collaboration with Penelope.ai
